# Inactivated virus vaccine BBV152/Covaxin elicits robust cellular immune memory to SARS-CoV-2 and variants of concern

**DOI:** 10.1101/2021.11.14.21266294

**Authors:** Rajesh Vikkurthi, Asgar Ansari, Anupama R Pai, Someshwar Nath Jha, Shilpa Sachan, Suvechchha Pandit, Bhushan Nikam, Anurag Kalia, Bimal Prasad Jit, Hilal Ahmad Parray, Savita Singh, Pallavi Kshetrapal, Nitya Wadhwa, Tripti Shrivastava, Poonam Coshic, Suresh Kumar, Pragya Sharma, Nandini Sharma, Juhi Taneja, Anil K Pandey, Ashok Sharma, Ramachandran Thiruvengadam, Alba Grifoni, Shinjini Bhatnagar, Daniela Weiskopf, Alessandro Sette, Pramod Kumar Garg, Nimesh Gupta

## Abstract

The characteristics of immune memory established in response to inactivated SARS-CoV-2 vaccines remains unclear. We determined the magnitude, quality and persistence of cellular and humoral memory responses up to 6 months after vaccination with BBV152/Covaxin. Here, we show that the quantity of vaccine-induced spike- and nucleoprotein-antibodies is comparable to that following natural infection and the antibodies are detectable up to 6 months. The RBD-specific antibodies decline in the range of 3 to 10-fold against the SARS-CoV-2 variants in the order of alpha (B.1.1.7) > delta (B.1.617.2) > beta (B.1.351), with no observed impact of gamma (P.1) and kappa (B.1.617.1) variant. We found that the vaccine induces memory B cells, similar to natural infection, which are impacted by virus variants in the same order as antibodies. The vaccine further induced antigen-specific functionally potent multi-cytokine expressing CD4^+^ T cells in ∼85% of the subjects, targeting spike and nucleoprotein of SARS-CoV-2. Marginal ∼1.3 fold-reduction was observed in vaccine-induced CD4^+^ T cells against the beta variant, with no significant impact of the alpha and the delta variants. The antigen-specific CD4^+^ T cells were populated in the central memory compartment and persisted up to 6 months of vaccination. Importantly the vaccine generated Tfh cells that are endowed with B cell help potential, similar to the Tfh cells induced after natural infection. Altogether, these findings establish that the inactivated virus vaccine BBV152 induces robust immune memory to SARS-CoV-2 and variants of concern, which persist for at least 6 months after vaccination. This study provides insight into the attributes of BBV152-elicited immune memory, and has implication for future vaccine development, guidance for use of inactivated virus vaccine, and booster immunization.

## Main Text

SARS-CoV-2 vaccines received emergency approval in several countries and are highly effective in reducing the disease burden due to COVID-19 pandemic. BBV152 is a whole-virion inactivated SARS-CoV-2 vaccine based on Asp614Gly variant and formulated with a toll-like receptor (TLR) 7/8 agonist molecule (imidazoquinolin) adsorbed to alum^1^. BBV152 is the first alum-imidazoquinolin adjuvanted vaccine approved for use in a large population. The vaccine is administered as two intramuscular doses of 6 µg inactivated virus 4-weeks apart. Almost everyone seroconverted after 4-weeks of complete vaccination^2^. Recently reported phase 3 data indicated an efficacy up to 78% against symptomatic infection, 93.4% from severe disease and 63.6% from asymptomatic disease^3^. Despite the global distribution and potential in providing clinically significant protection, limited evidence is available on the mechanism of immunity and the traits of immune memory established by BBV152.

The immune memory established after SARS-CoV-2 infection is expected to stay for a long-term. Many studies including ours have shown that memory T and B cells persist for several months after recovery from the symptomatic infection^4-7^. In fact, T cells are implicated in the less severe outcome of COVID-19 and mediate protection against SARS-CoV-2^8-10^. Moreover, T-cell memory after recovery from COVID-19 is mainly skewed towards the CD4^+^ T cells^4,11,12^. CD4^+^ T cells might be contributing to the protection against SARS-CoV-2 by limiting viral replication and dissemination, as well as by establishing good quality B cell and antibody responses in germinal centers. In fact, CD4^+^ T cells and B cells are detected for a long time even during rapidly waning levels of antibodies^13,14^. Thus, in addition to the antibody response, the cellular responses play an important role in conferring the long-term protective efficacy to a vaccine.

Studies largely carried out with mRNA vaccination highlight that protection may require low level of neutralizing antibodies along with other immune effector mechanisms including T cells and non-neutralizing antibodies^15,16^. Indeed, despite the decline in antibody response, infection with virus variants often lead to mild or asymptomatic disease after complete vaccination^17-19^ which further highlight the protective contribution of cellular responses. Considering that BBV152 provides an opportunity of exposing the whole virion to the human immune system and it is formulated with a first-in-human adjuvant, there is a growing interest to understand the traits of immune memory established by this vaccine and its effectiveness against the virus variants. The detailed knowledge on the humoral and cellular memory will be very helpful in guiding the wider use of this vaccine in pandemic control and deciding booster immunization.

Here, we investigated some of these key questions in 71 SARS-CoV-2 unexposed individuals who had received two doses of alum-imidazoquinolin adjuvanted BBV152 vaccine, up to 6 months after complete vaccination (Fig. 1a and Table 1). In order to understand the extent of humoral and cellular memory responses induced by BBV152, we further compared the vaccine-induced responses with the immune memory in 73 individuals recovered from mild COVID-19. Of note, the samples in infection group were collected between November-2020 to January-2021, prior to the Delta variant surge in India. We first measured anti-spike antibodies in the plasma samples from vaccinated and recovered individuals. All vaccinated and recovered individuals had detectable anti-spike IgG with a geometric mean end-point titer of 2×10^3^ (95% CI, 1.4×10^3^-3.7×10^3^) and 3×10^3^ (95% CI, 1.6×10^3^-5.8×10^3^) respectively (Fig. 1b). The anti-spike IgG titer was not significantly different between vaccination and post-recovery from infection. As the inactivated virus vaccine exposes multiple proteins of virus to the immune system, it is likely that proteins other than virus-spike will also be targeted in the vaccine-induced antibody responses. Like anti-spike IgG, we found that vaccine was capable of inducing anti-nucleoprotein IgG with no significant difference than the levels following natural infection (Fig. 1c). This observation is in line with the clinical trial data of BBV152 ^1^. We further examined the neutralizing potential of vaccine induced antibodies using the surrogate virus neutralization assay (sVNT), which relies on the presence of spike-ACE2 interaction inhibiting antibodies ^20^. The 1:10 dilution of plasma samples was examined in the sVNT assay. The antibodies showed neutralization potential in ∼92% (22/24) of the tested samples from vaccine group as compared to those with natural infection where all tested samples showed antibodies with neutralization ability (Fig. 1d). The virus neutralizing ability of antibodies was not different between vaccination and infection. Moreover, significant correlation between spike and nucleoprotein antibodies was found in vaccinated individuals suggesting that the vaccine induced coordinated immune response to different virus proteins, as observed in case of infection (Fig. 1 e-f). SARS-CoV-2 mRNA vaccines were shown to induce antibodies higher than the convalescence samples ^21^, which seems not to be the case with BBV152. However, BBV152 has an added advantage of inducing antibodies to other proteins that may be helpful in reducing cell-to-cell virus spread via effector mechanism ^22^. Moreover, like natural infection, the anti-spike and anti-nucleoprotein IgG were detectable up to at least 6 months after the complete vaccination (Extended data Fig.1 a-d). The presence of high titer IgG up to at least 6 months is very encouraging, as the vaccine-induced antibodies will be available in circulation in sufficient quantity to provide protective immunity. RBD binding antibodies directly correlate with the levels of virus neutralizing antibodies ^23^. Thus, to examine the broad protective potential of antibodies, we examined the efficacy of vaccine-induced antibodies against RBD protein of SARS-CoV-2 and its variants - alpha (B.1.1.7), beta (B.1.351), gamma (P.1), kappa (B.1.617.1) and delta (B.1.617.2). Like sVNT assay, ∼93% (44/47) of the tested subjects showed the presence of anti-RBD IgG. The vaccine-induced anti-RBD IgG was not affected by the gamma and kappa variant, however, the levels were significantly lower in case of the alpha, beta and delta variants, than that of the wild-type RBD (Fig. 1g). Most striking decline of 10-fold was observed in case of beta (34/47; P<0.0001) followed by the 7-fold decline against delta (42/47; P=0.002) and 3-fold decline to alpha variant RBD (43/47; P=0.03). Similar level of decline was observed in case of anti-RBD IgG from natural infection, except the decline was modest against delta variant (Fig. 1g). The highest decline against beta followed by delta and alpha variant was also observed in case of mRNA and the AstraZeneca vaccines ^24^. Certainly, among the recently circulating variants, beta (B.1.351) variant shows highest immune escape ability against COVID-19 vaccines or memory from infection. Surprisingly, as compared to BBV152, infection acquired anti-RBD antibodies were not impacted by the delta (B.1.617.2) and other variants. It’s plausible that the antibody maturation happens over time after the recovery from infection and that results in this increased neutralization potency and breadth ^7,25^. Unlike natural infection, the RBD antibody evolution does not happen over a long period of time in case of mRNA vaccines and most of the antibodies after 5 months of vaccination represent the antibodies that are populated during initial responses ^26^. Whether similar phenomena of limited antibody evolution over time exist in case of TLR7/8 agonist adjuvanted BBV152 vaccine, needs further investigation. Altogether, antibody analyses clearly establish that BBV152 elicits potent antibodies against virus-spike and -nucleoprotein that are capable of restricting the virus entry and further dissemination, and persist in high titers for at least up to 6 months. Moreover, the variant impact analyses suggest that BBV152 induced antibodies robustly respond against gamma and kappa variant. Although, a decline in anti-RBD IgG reactivity observed in the order of alpha>delta>beta variants, majority of the subjects still show the detectable antibodies that may contribute in the protection against these VOCs.

**Table 1.** Characteristics of participants included in the study.

| **Characteristics** | **BBV152**<br>**(Covaxin)** | **COVID-19**<br>**(Mild)** |
| --- | --- | --- |
| No. of participants | **n=71** | **n= 73** |
| Male | 50 (70.4%) | 47 (64.4%) |
| Female | 21 (29.6%) | 26 (35.6%) |
| Age, in years Median (IQR) | 43 (18) | 33 (21) |
| Confirmed SARS-CoV-2 infection (RT-PCR) | 0 | 73 (100%) |
| Sample collection post vaccination/post diagnosis, in days Median (IQR) | 115 (45) | 224 (56.3) |
| Duration from first to second dose, in days Median (IQR) | 29 (3) | -- |
| **Comorbidities** |  |  |
| Cardiovascular diseases | 1/71 (1.4%) | 3/73 (4.1%) |
| Diabetes | 6/71 (8.5%) | 6/73 (8.2%) |
| Hypertension | 3/71 (4.2%) | 9/73 (12.3%) |
| Cancer | 0 | 0 |
| Chronic respiratory diseases | 0 | 0 |
| **Symptoms in COVID-19 patients** |  |  |
| Fever | 42/73 (57.5%) |  |
| Cough | 29/73 (39.7%) |  |
| Headache | 14/73 (19.2%) |  |
| Sore throat | 30/73 (41.1%) |  |
| Body ache | 22/73 (30.1%) |  |
| Loss of smell | 18/73 (24.7%) |  |
| Loss of taste | 19/73 (26%) |  |
| Vomiting | 7/73 (9.6%) |  |
| Nausea | 7/73 (9.6%) |  |
| Abdominal pain | 4/73 (5.5%) |  |
| Diarrhoea | 9/73 (12.3%) |  |
| Breathlessness | 13/73 (17.8%) |  |
| Runny nose | 8/73 (11%) |  |
| Haemoptysis | 2/73 (2.7%) |  |
| Fatigue | 16/73 (21.9%) |  |

**Fig. 1.**
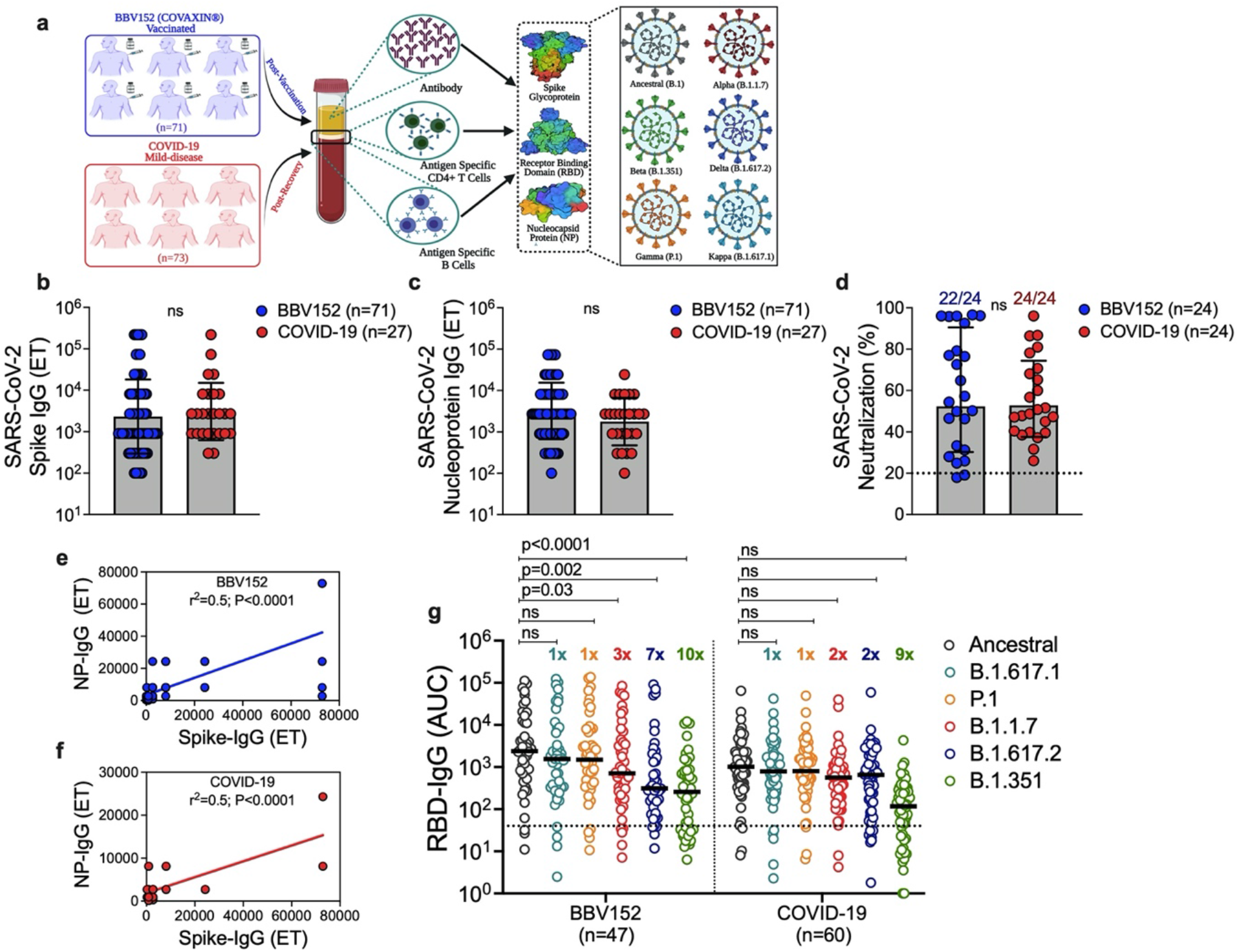
BBV152/Covaxin induces robust antibody response to SARS-CoV-2 and its variants. (**a**) Study design to investigate the BBV152-elicited immune memory against SARS-CoV-2 and its variants. The end-point titer of SARS-CoV-2 specific antibodies measured by in-house ELISA in plasma samples derived from BBV152/Covaxin vaccinated individuals (“BBV152”) and individuals recovered from mild COVID-19 (“COVID-19”) for (**b**) Spike protein (**c**) Nucleoprotein. (**d**) neutralization potential of antibodies measured by the SARS-CoV-2 surrogate virus neutralization test in vaccinated and recovered individuals. Correlation between anti-spike and anti-nucleoprotein IgG in (**e**) vaccinated and (**f**) COVID-19 subjects. (**g**) Comparison of anti-RBD IgG levels measured as area under the curve (AUC) in response to ancestral SARS-CoV-2 and its variants; alpha (B.1.1.7), beta (B.1.351), gamma (P.1), kappa (B.1.617.1) and delta (B.1.617.2) in vaccinated and mild COVID-19 recovered individuals. The fold reduction in IgG levels to variant-RBD with respect to ancestral SARS-CoV-2-RBD is represented at the top of dataset for each variant. Black bars indicate geometrical mean and geometric SD in (**b-d**) and median in (**g**). Dotted line represents the cut-off for positivity. Statistics by (**b-d**) two-tail Mann-Whitney test, (**e-f**) Pearson’s correlation coefficient, (**g**) mixed-effect analysis followed by Tukey’s multiple comparisons. ns: non-significant.

B cell memory is a critical component in durability of protection conferred by vaccine. We thus examined the memory B cells established in response to the BBV152. To examine the phenotype of SARS-CoV-2-specific B cells, we utilized R848 and IL2 stimulation for polyclonal activation and differentiation of memory B cells in to antibody secreting cells (ASCs). The RBD-specific ASCs were then determined using the ELIspot assay (Fig. 2a). Majority of the subjects showed the presence of circulating SARS-CoV-2 specific memory B cells, both in vaccination and infection (Fig. 2 b-d). The vaccine-induced IgG^+^ B cells represent around 0.44±0.1% (median, 0.13%) of the total IgG^+^ cells, which was in the comparable range of IgG^+^ B cells established after the recovery from mild COVID-19 0.62±0.2% (median, 0.31%) (Fig. 2b). Like natural infection (0.13±0.02%, median, 0.11), the vaccine-induced IgA^+^ B cells were also present in the similar frequency (0.24±0.05%, median, 0.17) (Fig. 2c). The proportion of IgM^+^ cells was higher in the range of 1.3±0.16% of the total IgM^+^ B cells (Fig. 2d). Our finding in case of recovered individuals corroborate with the previous work that showed similar range of IgG^+^ and IgA^+^ B cells after recovery from mild infection ^14^. These results also establish that BBV152 induces memory B cells in the similar proportion like natural infection. With a probable decline in antibodies over time, memory B cells will be most likely present to initiate an immediate recall response to virus, or to a vaccine booster. We then determined the breadth of RBD-specific B cells in their reactivity to the recently circulating SARS-CoV-2 variants (Extended data Fig. 2 a-b). The vaccine-induced IgG^+^ B cells showed no significant decrease in response to the kappa and gamma variant (Fig. 2e). The modest reduction of 1.5 to 2-fold was observed against the alpha (B.1.1.7) and beta (B.1.351), over the RBD-specific IgG^+^ B cells established against the ancestral virus (Fig. 2e). Moreover, in a subset of samples, ∼2-fold reduction in vaccine-induced IgG^+^ B cells was observed against the delta (B.1.617.2) variant (Extended data Fig. 2c). Interestingly, no significant impact on IgA^+^ or IgM^+^ B cells was observed due to any of the variants tested (Fig. 2 f-g; Extended data Fig. 2 d-e). IgG^+^ B cells established after the mild infection showed similar trend in their reactivity towards the SARS-CoV-2 variants, except, higher decrease was observed against the alpha, beta and delta variants (Fig. 2h; Extended data Fig. 2f). Like BBV152, no significant impact of variants was observed in the infection-acquired IgA^+^ and IgM^+^ B cells (Fig. 2 i-j; Extended data Fig. 2 g-h). Clearly, BBV152 induced memory B cells were mostly sustained against the VOCs as compared to the circulating memory B cells acquired after the recovery from natural infection (Fig. 2k). Whether the adjuvant used in BBV152 formulation is contributing to this larger breadth in memory B cells remains to be defined. No impact of VOCs on the BBV152- or infection-induced IgA^+^ B cells is encouraging. Dimeric IgA antibodies are more potent than IgG in neutralizing SARS-CoV-2 ^27^ and IgA^+^ B cells with mucosal homing traits were found in SARS-CoV-2 infection ^28^. Thus, for a broadly directed breadth of an effective protection, it’ll be worthwhile to determine whether IgA^+^ B cells established by BBV152 localized and persist in the upper respiratory tract.

**Fig. 2.**
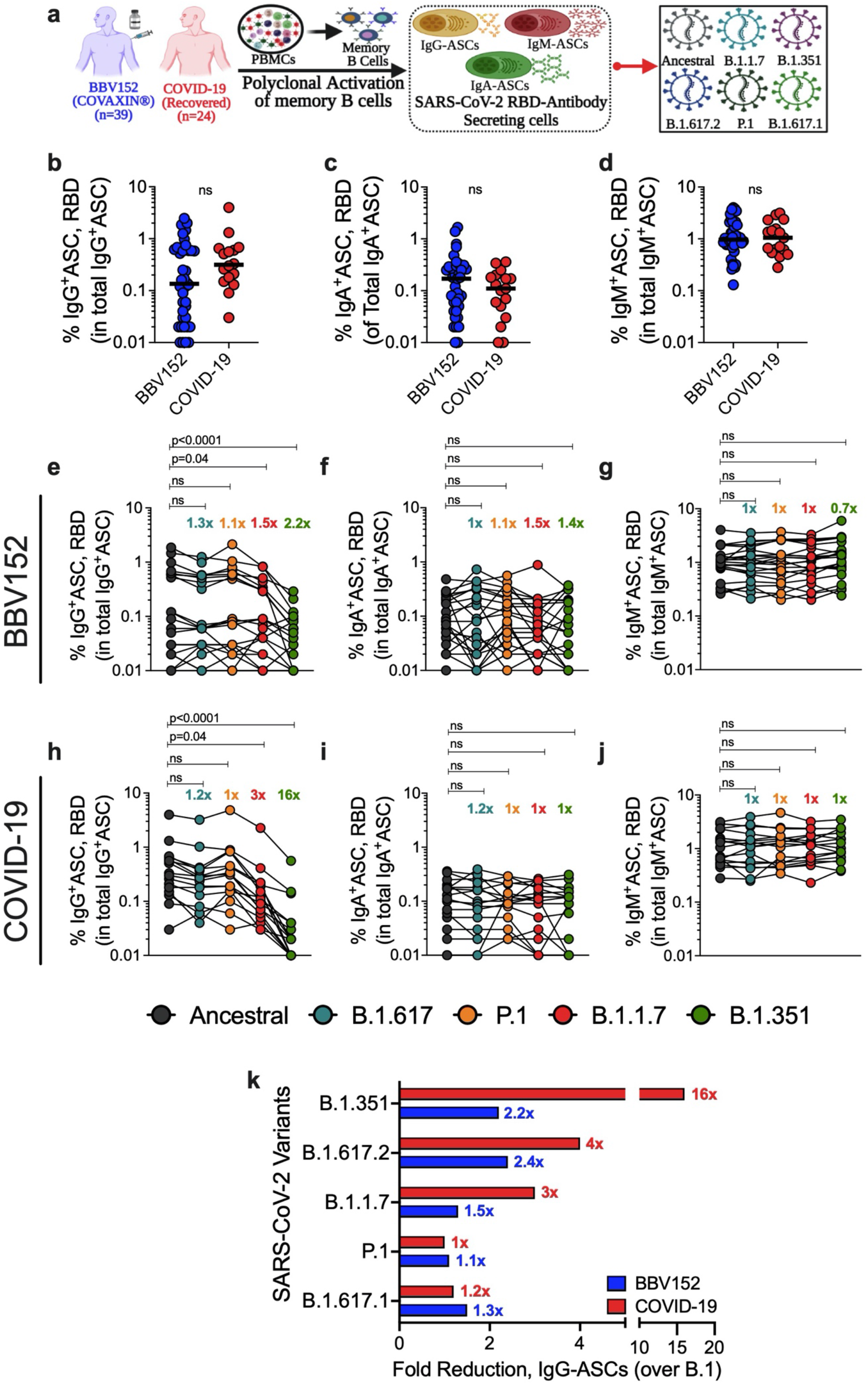
BBV152/Covaxin generate memory B cells with potent cross-reactivity to SARS-CoV-2 variants. (**a**) Experimental design for measuring SARS-CoV-2-specific memory B cells and their isotype in Fluorospot assay utilizing the PBMCs from vaccinated (BBV152, n=39) and recovered individuals (COVID-19, n=24). Proportion of SARS-CoV-2-RBD-specific antibody secreting cells (ASCs) calculated in the total corresponding isotype of ASCs in 1 million PBMCs for (**b**) IgG^+^ ASCs (**c**) IgA^+^ ASCs and (**d**) IgM^+^ ASCs, in vaccination and infection. Black bars indicate median. Comparison of the frequency of ancestral SARS-CoV-2 RBD-specific memory B cells and their reactivity to RBD protein of variants; alpha (B.1.1.7), beta (B.1.351), gamma (P.1), kappa (B.1.617.1) and delta (B.1.617.2) for (**e**) IgG^+^ ASCs (**f**) IgA^+^ ASCs and (**g**) IgM^+^ ASCs in BBV152 vaccination, and for (**h**) IgG^+^ ASCs (**i**) IgA^+^ ASCs and (**j**) IgM^+^ ASCs in recovery from mild COVID-19. Fold reduction in RBD-reactive B cells against variants over the RBD-specific B cells to ancestral virus is depicted in top of graph for each variant. (**k**) Comparison of the quantitative reduction in IgG^+^ memory B cells against multiple variants between vaccination and natural infection. Statistics by (**b-d**) two-tail Mann-Whitney test, (**e-j**) one way ANOVA followed by Dunnett’s multiple comparisons. ns: non-significant.

By killing infected cells and by supporting B cell function and antibody response, T cells are vital mediators in the protective response to SARS-CoV-2. We thus investigated the SARS-CoV-2 specific memory CD4^+^ T cells induced in response to the BBV152 vaccination. To detect antigen-specific T cells, we performed the flow cytometry activation-induced marker (AIM) assay utilizing OX40/CD137 activation markers (Extended Fig. 3a) and overlapping peptides megapool (MP) spanning the SARS-CoV-2 antigens of ancestral virus and VOCs ^11,29^. We also compared the reactivity of CD4^+^ T cells to SARS-CoV-2 and its variants in individuals who either received BBV152 or recovered from infection (Fig. 3a-b). The magnitude of total CD4^+^ T cells was not different in the vaccinated or recovered individuals (Fig. 3c). BBV152 induced a robust spike-specific CD4^+^ T cell response (0.01 to 1.5%; median, 0.18%), which was of similar magnitude as observed in recovery from mild infection (0.01 to 1.35%; median, 0.29) (Fig. 3d). The Fisher’s analysis on the T-cell response in unstimulated (DMSO) vs spike peptide_MP_ stimulated conditions suggested that spike-specific T cells were present in detectable levels in ∼85% of the vaccinated (38/45) and ∼90% of the infected individuals (36/40) (Fig. 3d). In interferon-γ (IFN-γ) Fluorospot assay, we confirmed that the vaccine-induced spike-specific T cells were capable of effector function in secreting IFN-γ during the antigen recall response (IFN-γ SFC, DMSO: 22±8; Spike: 98±21; P=0.002) (Extended Fig. 4 a-b). We further analyzed the levels of T-cell specific cytokines in culture supernatant of PBMCs stimulated with spike peptide_MP_. The cytokine levels were not significantly different between BBV152 and COVID-19 convalescent samples, suggesting comparable effector functions of T cells in vaccination and infection (Extended Fig. 5 a-f). Upon antigen stimulation, the cells produced highest amount of TNF-α (mean concentration (pg/ml)) (BBV152: 3413; COVID-19: 4341) followed by IL2 (BBV152: 113; COVID-19: 88), IFN-γ(BBV152: 32; COVID-19: 9) and IL17 (BBV152: 26; COVID-19: 24), with a minimal contribution of IL13 (BBV152: 18; COVID-19: 6) and IL4 (BBV152: 5; COVID-19: 5) (Extended Fig. 5g). Clearly, like infection, a Th1-skewed multifunctional profile was observed for CD4^+^ T cells established after BBV152 vaccination. It’s a notable outcome of BBV152, because the Th1-cell skewed response and IFN-γ secreting T cells are associated with better protection from the disease ^30,31^. Previously, TLR7/8 agonist was shown to elicits multifunctional T cell responses in non-human primates^32^. It’s likely that TLR7/8 agonists adjuvant in BBV152 formulation mediated this potent effector function to the vaccine-induced CD4^+^ T cells. Similar profile of multifunctional T cells secreting high TNF-α>IL2>IFN-γ are also seen in SARS-CoV-2 mRNA vaccination and adenoviral-vectored vaccines ^21,33,34^. Further investigation in this line will be helpful to establish if multifunctional T cells can be used as the cellular correlates of protection for COVID-19 vaccines, as described in case of Leishmania infection ^35^. In addition to spike, BBV152 provides an opportunity to direct T cells against other proteins of the SARS-CoV-2.

**Fig. 3.**
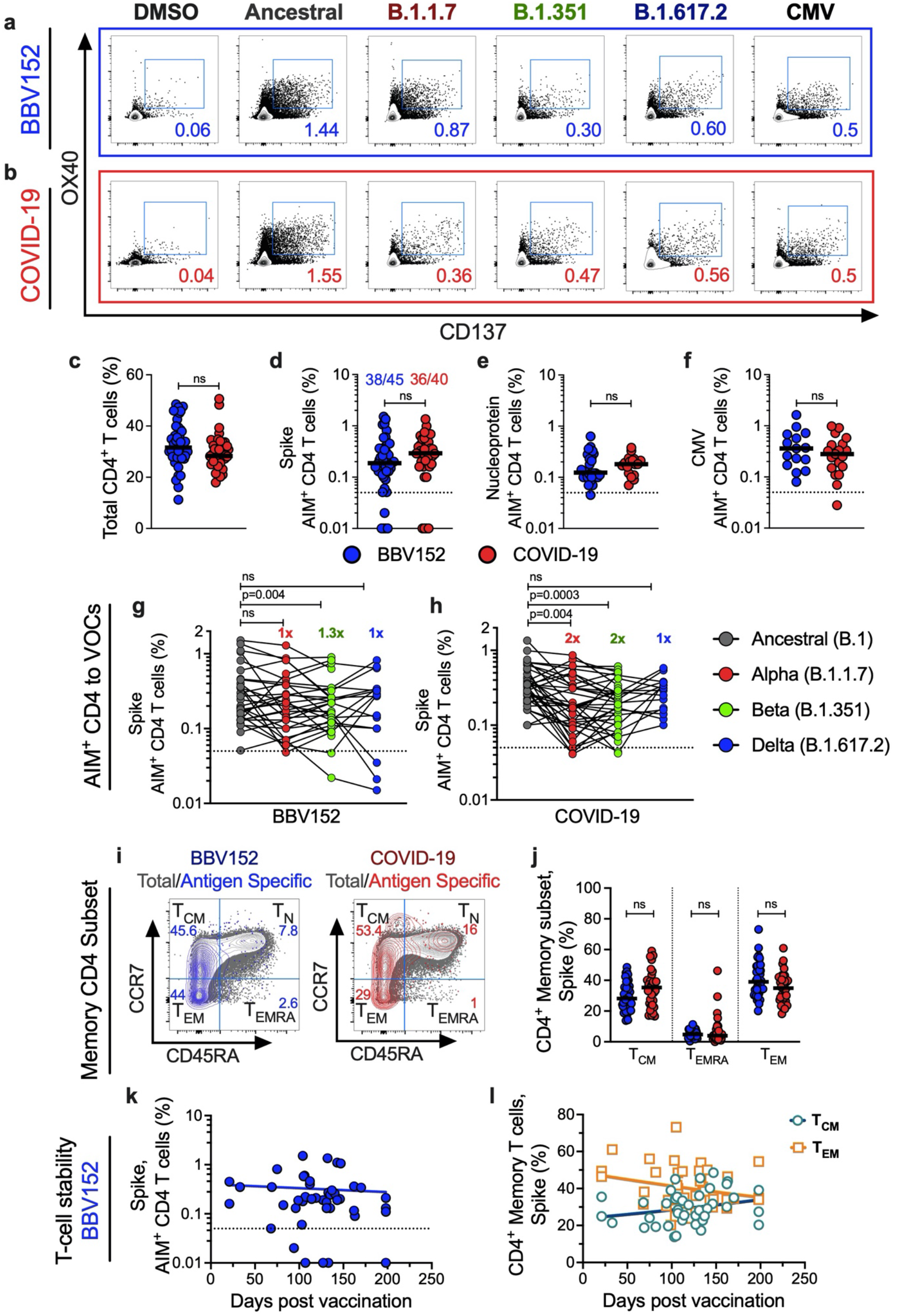
BBV152 vaccination induces SARS-CoV-2 specific memory CD4^+^ T cells that robustly respond to major variants of concern. SARS-CoV-2-specific memory CD4^+^ T cells were quantified in PBMCs derived from the individuals vaccinated with BBV152 (“BBV152”) and individuals recovered from mild COVID-19 (“COVID-19”). PBMCs were stimulated with overlapping peptide megapools (MP) spanning spike glycoprotein of ancestral SARS-CoV-2, nucleoprotein of SARS-CoV-2 or the MP specific to the spike protein of VOCs; alpha (B.1.1.7), beta (B.1.351), and delta (B.1.617.2). Antigen-specific cells (OX40^+^CD137^+^) were determined after 24h stimulation of PBMCs in the AIM assay. Equimolar DMSO was used as the negative control and CMV MP was used as the positive stimulation control. Representative gating strategy for measuring the antigen-specific AIM^+^ (OX40^+^CD137^+^) CD4^+^ T cells in the total CD4^+^ T cells from (**a**) BBV152 and (**b**) COVID-19 (see extended Fig. 3 for detailed gating strategy). (**c**) Frequency of total CD4^+^ T cells in BBV152 and COVID-19, (**d**) Comparison of background subtracted spike-specific CD4^+^ T cells in BBV152 and COVID-19 groups, (**e**) Comparison of background subtracted nucleoprotein-specific CD4^+^ T cells in BBV152 and COVID-19, (**f**) CMV-specific CD4^+^ T cells in BBV152 versus COVID-19. Quantitation of spike-specific CD4^+^ T cells responding to ancestral SARS-CoV-2 and VOCs - B.1.1.7, B.1.351 and B.1.617.2 in individuals after (**g**) vaccination and (**h**) recovery from COVID-19. Values in parentheses represents fold-reduction in spike-specific CD4^+^ T cells responding to VOCs over ancestral virus. (**i**) Representative FACS plots showing the memory subsets among SARS-CoV-2 spike-specific CD4^+^ T cells after BBV152 vaccination (blue) or recovery from COVID-19 (red), over total memory subsets (grey), determined as Central memory (T_CM_: CD45RA^-^ CCR7^+^), Effector memory (T_EM_: CD45RA^-^CCR7^-^), or terminally differentiated effector memory cells re-expressing CD45RA (T_EMRA_: CD45RA^+^CCR7^-^). (**j**) Comparison of the proportion of memory subsets in spike-specific CD4^+^ T cells between BBV152 and COVID-19. (**k**) Persistence of spike-specific CD4^+^ T cells up to 6 months after BBV152 vaccination, (**l**) Proportion of central (T_CM_) and effector (T_EM_) memory subsets in the persisted spike-specific memory CD4^+^ T cells after BBV152 vaccination. Black bars indicate median. Dotted line represents the limit of positivity. Statistics by (**c-f**) two-tail Mann-Whitney test, (**g-h**) Wilcoxon signed-rank test (**j**) 2way ANOVA followed by Tukey’s multiple comparisons, (**k-l**) non-linear regression. ns: non-significant.

After spike, nucleoprotein is the majorly targeted virus protein by infection induced CD4^+^ T cells ^36^. Thus, to determine the broadly directed T cells, we next examined the nucleoprotein-specific T cells in BBV152 vaccinated individuals. Like spike T cells, high frequency nucleoprotein-specific T cells were also induced in vaccinated individuals, which were comparable with the extent of nucleoprotein-specific T cells in mild infection (median, BBV152: 0.12%; COVID-19: 0.18%; P=ns) (Fig. 3e). This is encouraging as broadly directed T cells may supplement spike-specific T-cell response in the protection against SARS-CoV-2 and its variants. No significant difference was observed between vaccination and infection for T cells to an unrelated antigen, CMV (Fig. 3f).

We next measured the CD4^+^ T-cell response directed to spike protein of ancestral virus and the recently circulating variants of concern – alpha (B.1.1.7), beta (B.1.351) and delta (B.1.617.2). The BBV152 induced CD4^+^ T cells were not reduced significantly against alpha and delta spike protein as compared to the ancestral virus spike (mean %, Ancestral: 0.39±0.07; B.1.1.7: 0.31±0.05, B.1.617.2: 0.28±0.05) (Fig. 3g). Although a significant decline was observed in case of beta variant, the median reduction was 1.3-fold over the ancestral virus (B.1.351: 0.25±0.04; P=0.004). The reactivity profile of CD4^+^ T cells acquired from natural infection was comparable to the vaccine, except the median decline was around ∼2-fold for alpha (P=0.004) and beta (P=0.0003) (mean %, Ancestral: 0.39±0.04; B.1.1.7: 0.25±0.04; B.1.351: 0.24±0.02; B.1.617.2: 0.29±0.03) (Fig. 3h). Importantly, majority of the samples showed memory T cells in detectable limits even after the observed decline. We further assessed the memory phenotype of virus-specific CD4^+^ T cells in the individuals with positive T cell response to the vaccine. The gating strategy is shown in the extended Figure 3a, and in Fig. 3i where AIM^+^ cells are depicted as the antigen-specific cells in case of both vaccine and infection. Like infection, vaccine-induced AIM^+^ T cells were also enriched in both the central (CD45RA^-^CCR7^+^) and effector memory (CD45RA^-^CCR7^-^) compartments (Fig. 3j). The enrichment of antigen-specific CD4^+^ T cells in the central and effector memory compartment is significant, as the memory CD4^+^ T cells are required at the time of recall response against SARS-CoV-2 and its variants. Importantly, vaccine-induced SARS-CoV-2 specific CD4^+^ T cells persisted at least up to 6 months (Fig. 3k) with both the central and effector memory T cells enriched in these persisted CD4^+^ T cells (Fig. 3l). It seems that vaccine-induced central memory T cells are more durable than the effector memory subset, which is an obvious biological process in the differentiation of memory CD4^+^ T cells ^37^.

Tfh cells are essential for the generation of optimal quality humoral responses and thus, they are vital subset of CD4^+^ T cells required for the protective immunity against pathogens ^38^. Tfh-cell response is reported in both SARS-CoV-2 infection ^9,39^ as well as in mRNA vaccination ^33,40^. We thus examined the potential of BBV152 in inducing the Tfh cells. Frequency of antigen-specific Tfh cells for spike (Fig. 4a-b) and nucleoprotein (Fig. 4c-d) were measured by gating the CXCR5^+^PD1^+/-^ cells in the AIM^+^ antigen-specific CD4^+^ T cells. Spike specific Tfh cells were present in the range of 0.03% to 0.43% of total CD4^+^ T cells in the vaccinated individuals (median, 0.11%), which was marginally lower than the Tfh cells acquired after recovery from mild infection (0.06% to 0.38%, median, 0.15%; P=0.03) (Fig. 4b). Tfh cells specific to nucleoprotein were also induced in response to the vaccine (0.03% to 0.29%; median, 0.07%), with the comparable extent like infection (0.06% to 0.26%; median, 0.09%; ns) (Fig. 4d). Our observation is in line with previous report where individuals with mild disease favor more efficient Tfh responses ^41^. The duration of antigen persistence is a critical driver in the generation of Tfh cells ^42^. In fact, active germinal centers for 3 to 4 months are reported in individuals after infection ^43^. Thus, it’s possible that the lower magnitude of spike-Tfh cells in case of vaccination is due to an inferior persistence of inactivated virus antigens leading to shorter duration of germinal center response as compared to the mild-infection. Circulating Tfh (cTfh) cells are divided depending on the surface expression of CCR6 and CXCR3 in to Tfh1 (CXCR3^+^CCR6^-^), Tfh2 (CXCR3^-^CCR6^-^) or Tfh17 (CXCR3^-^CCR6^+^) subsets (Extended Fig. 6a) ^44^. BBV152 induced cTfh cells seem to comprise of all three subsets with the highest enrichment of Tfh2 and Tfh1 cells (Extended Fig. 6b). Similar heterogeneity was observed in case of cTfh cells acquired after infection (Extended Fig. 6b). Unlike spike-Tfh cells, nucleoprotein-Tfh cells showed preferential enrichment in Tfh2 polarized subset for both vaccination and infection (Extended Fig. 6c). Functional relevance of cTfh heterogeneity is not clearly defined and it may vary in different pathological conditions ^45,46^. It remains to be defined how Tfh heterogeneity against spike or nucleoprotein is implicated in the vaccine-induced protective immune responses.

**Fig. 4.**
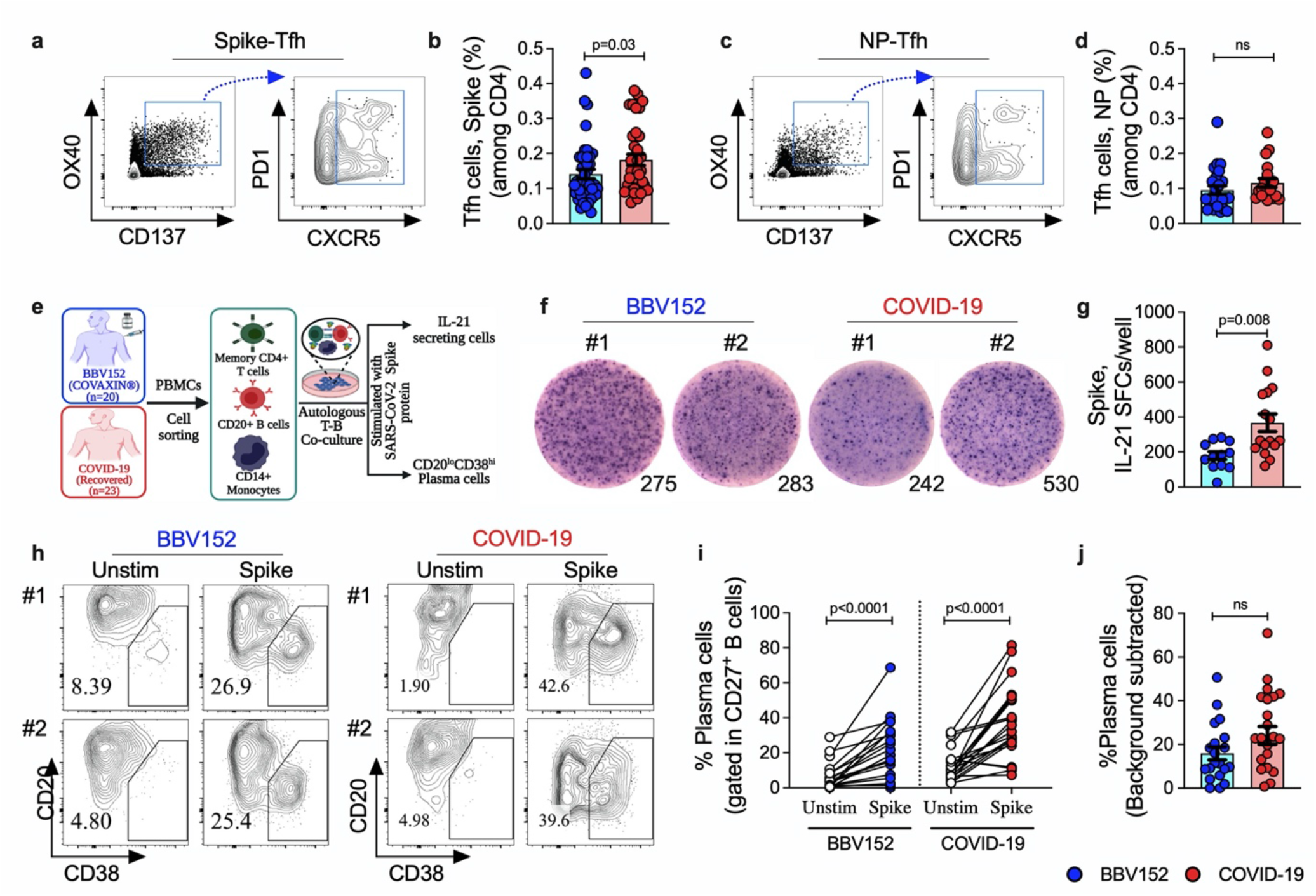
BBV152 vaccination establish SARS-CoV-2 specific follicular T helper (Tfh) cells that are functionally potent in inducing humoral immunity. (**a**) Representative FACS plot depicting the gating strategy for quantitation of circulating Tfh (CXCR5^+^PD1^+^) cells in spike-specific CD4^+^ T cells, (**b**) Comparison of spike-specific circulating Tfh cells frequency in total CD4^+^ T cells after BBV152 vaccination or recovery from mild COVID-19. Similar analyses for nucleoprotein-specific Tfh cells showing (**c**) Representative FACS plots and (**d**) Comparison of nucleoprotein-specific Tfh cells in BBV152 and COVID-19 groups. (**e**) Experimental design for the qualitative assessment of SARS-CoV-2 spike-specific memory CD4^+^ T cells for their potential in inducing humoral immunity, as determined by enumeration of IL-21 secreting cells and generation of plasma cells in autologous T-B co-cultures stimulated with full length spike protein for 9 days. (**f**) Representative images of IL21 ELIspot depicting IL-21 spot forming cells (SFCs)/well, in two of the representative donors from BBV152 vaccination and COVID-19, (**g**) Comparison of the magnitude of IL-21 secreting cells between BBV152 and COVID-19. (**h**) FACS plots representing the frequency of CD20^low^CD38^hi^ plasma cells gated on CD27^+^ memory B cells in unstimulated (Unstim) or spike protein (spike) stimulation, in two donors each from BBV152 and COVID-19 groups, (**i**) magnitude of plasma cells (as the percentage of CD27^**+**^ B cells) in unstimulated and spike-stimulated conditions for BBV152 and COVID-19, (**j**) Comparison of the frequency of background-subtracted spike-specific plasma cells between BBV152 vaccination and recovery from mild COVID-19. Black bars indicate mean±s.e.m. Statistics by (**b, d, g, j**) two-tail Mann-Whitney test, (**i**) Wilcoxon signed-rank test. ns: non-significant.

The functional assessment of CD4^+^ T cells in providing help to B cells may serve as an important immunological parameter for qualitative evaluation T cells in infection or vaccination. The B-cell help potential of CD4^+^ T cells is not yet demonstrated in COVID-19 infection or vaccination. We recently described that supplementation of conventional T-B co-cultures with autologous monocytes substantially enhance the sensitivity and efficiency in defining the B-cell help potential of SARS-CoV-2 specific memory CD4^+^ T cells ^47^. We thus utilized the newly described method to examine the help potential of BBV152-induced CD4^+^ T cells by enumeration of IL-21 secreting cells and measuring the extent of plasma cells differentiation in the autologous T-B co-cultures (Fig. 4e). Total B cells and memory CD4^+^ T cells were sorted from the PBMCs of vaccinated and recovered individuals and co-cultured for 9 days, along with autologous CD14^+^ monocytes and stimulated with full-length spike protein of SARS-CoV-2 (Fig. 4e). Like spike-Tfh, high frequency of IL21 secreting cells were present in recovered individuals than the vaccinated subjects (COVID-19: 367±50; BBV152: 179±22; P=0.008) (Fig. 4f-g). The plasma cell output reflects the threshold of help signals received from T cells. We thus determined the plasma cell output in these co-cultures by measuring the frequency of CD20^lo^CD38^hi^ plasma cells in CD3^-^CD19^+^CD27^+^ memory B cells (Fig. 4h). As expected, plasma cell output was significantly higher in spike-stimulated conditions over unstimulated conditions (Fig. 4i). Clearly, CD4^+^ T cells from vaccinated individuals were equally capable of inducing the plasma cell output like the T cells from recovered individuals (Fig. 4j) (BBV152: 16±3%; COVID-19: 24±4%; ns). These data suggest that the vaccine is capable of generating adequate quality CD4^+^ T cells with potent help functions like that acquired following natural infection.

## Discussion

The BBV152 vaccine has the dual advantage of inclusion of whole virus as well as the utilization of new adjuvant, alum-adsorbed imidazoquinolin. It is important to understand the mechanism of protective immunity and the durability of immune memory conferred by this vaccine. Understanding the traits of immune memory established by this vaccine will provide relevant knowledge on future vaccine development and the application of TLR7/8 agonist molecules as an adjuvant. Here, studying 71 subjects after complete vaccination up to 6 months, we found that BBV152 induced a robust immune memory against spike and nucleoprotein that was comparable to that following SARS-CoV-2 infection for the levels of antibodies, memory B cells, and memory CD4^+^ T cells. As demonstrated here, concurrent antibody and cellular memory responses are generated against spike and other proteins like nucleoprotein of SARS-CoV-2. Almost all of the current VOCs show major mutations in the spike protein of virus. The immune memory against conserved nucleoprotein may provide an added advantage over spike-only responses in protection against immune escape variants. Moreover, BBV152 is inducing a decent quantity of memory B cells that comprises of cells with all three isotypes IgG, IgA and IgM. It is assuring that even after natural decline in antibodies, memory B cells will provide restoration of the antibodies during secondary virus exposure. It is likely that a direct impact of adjuvant on the B cell compartment is contributing to this potential of BBV152.

We found spike-specific memory CD4^+^ T cells in the majority (∼85%) of the subjects after complete vaccination that comprised of both the central and effector memory T cells, and persisted for at least up to 6 months. The quantity of antigen-specific memory T cells comparable to that of natural infection, and the composition of memory subsets is an indicative of a long-term durability of BBV152-induced T cell responses. Like natural infection, the vaccine-elicited T cells exhibited Th1-skewed anti-viral functional profile with the cells expressing multi-cytokines like TNF-α, IL2 and IFN-γ. These findings on the functionality of BBV152 induced memory T cells is highly encouraging. It clearly assures that the vaccine has potential to confer protection via T-cell governed effector mechanisms. The memory pool consists of a substantial fraction of Tfh cells specific to multiple virus proteins and comparable to that of natural infection. A sizable quantity of Tfh cells with demonstrated potential of B-cell help further suggest that the adjuvanted vaccine has the ability to induce robust long-term humoral immunity.

Vaccine-elicited memory responses stand well against the recent virus variants with the impact observed in the manner of alpha>delta>beta variants, mostly on the antibodies, followed by moderate impact on memory B cells and well-preserved T-cell responses. The kappa variant that circulated in India and the UK prior to the delta surge and the gamma variant originated from Brazil showed no impact on humoral responses elicited by BBV152. Certainly, the larger breadth of BBV152-elicited cellular responses in protection against the VOCs is in line with the immune memory acquired from natural infection. Modest decline in humoral response and well-preserved memory T cell responses clearly underline that the immune memory from previous SARS-CoV-2 exposure and BBV152 vaccination was substantially effective against the delta variant. Indeed, beta (B.1.351) variant that is considered to be the highest immune escape version of VOCs was also limited in escaping from the BBV152-induced T cell responses. These findings clearly establish that the vaccine-induced T cells are capable of mediating protective response to SARS-CoV-2 variants. The protective contribution of the vaccine-induced T cells is further supported by the phase 3 effectiveness data that shows clinically significant protection from the asymptomatic infection and significantly lower viral load in the delta positive vaccine recipients as compared to the placebo group ^3^. Thus, the data reported here provides an understanding on the immunological basis of vaccine effectiveness.

Limitations of this study includes a modest sample size and limited blood volume to perform simultaneous analyses of multiple cellular responses to SARS-CoV-2 and VOCs. Although, the analyses from 4 weeks post-2^nd^ dose up to 6 months suffice for key information on the persistence of immune memory, the durability of immune memory cannot be defined in the absence of analyses at the baseline and the longitudinal follow up. The detailed immunological investigation in the larger prospective cohort of vaccine effectiveness is required for identifying the immune correlates-of-protection and understanding the mechanism of protective immunity conferred by BBV152.

BBV152-induced protective immunity seems to comprise of a low magnitude of neutralizing antibodies with potent T-cell response and a larger breadth of cellular reactivity against variants. The effective presence of memory T cells and antibodies in the respiratory tract will be ideal to control not only the SARS-CoV-2 infection but also the disease transmission. Thus, it’ll be worth investigating if BBV152 booster immunization via intranasal route can mediate localization of the protective immune memory in the local sites of infection or upper respiratory tract. With the encouraging demonstration of the potential of BBV152 in generating immune memory and its effectiveness against the recent virus variants, our study provides an assuring foundation for the future application of BBV152, an adjuvanted inactivated virus vaccine, in pandemic control response.

## Methods

### Ethics Statement

The study protocols were approved by the Human Research Ethics Committees of National Institute of Immunology and the partnering institutions. All associated procedures were carried out in accordance with approved guidelines. All participants provided written informed consent before enrolment for the study.

### Subject Recruitment and Sample Collection

The vaccine cohort includes the healthy subjects who received full Covaxin/BBV152 vaccination. Individuals who reported SARS-CoV-2 positivity either by RT-PCR any time before blood collection or experienced COVID-19 related symptoms were excluded from the study. Another cohort of SARS-CoV-2 convalescent individuals was used to compare the vaccine-induced immune responses to the immune responses generated in case of natural infection. The individuals with confirmed diagnosis for COVID-19 by RT-PCR test were recruited in the infection cohort. At the time of infection, none of the patients required hospitalization and all patients experienced mild manifestation of the disease (WHO Criteria). The demographic details and characteristics of participants in both the vaccine and infection cohort are provided in Table 1. Whole blood was collected from vaccinated participants and patients recovered from mild COVID-19, in tubes containing K3EDTA as anticoagulant. Plasma was separated from blood samples by centrifugation at 20°C and stored at -80 °C until further use. PBMCs were isolated using Ficoll Paque Plus (GE Life Sciences) density gradient medium and cryopreserved in multiple aliquots in Fetal Bovine Serum (Gibco) containing 10% Dimethyl Sulfoxide (DMSO; Thermo-Fisher) and stored in liquid nitrogen until further use in the assays. For all the cell based assays, PBMCs were obtained with >80% viability as assessed by acridine orange and propidium iodide double staining in the LUNA-FL (Logos Biosystems Inc., USA) cell counter.

### Recombinant protein from SARS-CoV-2 and its variants

Recombinant RBD proteins of wild type as well as variants of SARS-CoV-2 were either procured from Native Antigen, UK or expressed and purified from mammalian expression system, as described previously ^48^. Briefly, the desirable mutations were introduced into the wild type (B.1) RBD construct through site directed mutagenesis one by one for the characteristics mutations in VOCs - Kappa (B.1.617.1; E484Q, L452R), Gamma (P.1; K417T, E484K, N501Y), Alpha (B.1.1.7; N501Y), Beta (B.1.351; K417N, E484K, N501Y) and Delta (B.1.617.2; L452R, T478K). The sequence verified plasmid DNA representing mutant as well as wild type RBD sequences was further transfected to Expi293 cells using ExpiFectamine 293 Transfection Kit (ThermoFisher), and the secreted protein was purified from the supernatant. Highly purified mammalian-system expressed full length Spike protein and Nucleoprotein of wild-type SARS-CoV-2 were procured from the Native Antigen, UK.

### Detection of SARS-CoV-2-specific antibodies

The plasma IgG level specific to SARS-CoV-2 proteins was measured by enzyme-linked immunosorbent assay (ELISA), as described ^4^. The IgG response was analysed against full length Spike protein, Nucleoprotein and RBD protein of wild-type and its variants; Alpha (B.1.1.7), Beta (B.1.351), Gamma (P.1), Kappa (B.1.617.1) and Delta (B.1.617.2). Briefly, ELISA microtiter plates (Nunc, Maxisorp) were coated with 100µl/well of respective SARS-CoV-2 antigens in PBS (pH 7.4) at the final concentration of 1µg/ml and incubated overnight at 4°C. After blocking with PBS containing 3% Skim milk and 0.05% Tween-20, serially diluted heat inactivated plasma samples were added and incubated for 1.5 hours at room temperature. Plates were washed before incubation with 1:4000 dilution of HRP-conjugated anti-human IgG (Southern Biotech) for 1 hour at room temperature. Plates were washed and developed using the OPD-substrate (Sigma-Aldrich), stopped using HCl and optical density (OD) was measured at 492 nm. The antigen coated wells that were added with sample diluent alone were used as the blank. The OD values obtained in test wells after subtracting the mean of blank OD values were used for calculating the end-point titres or the AUC (Area Under the Curve) with a baseline of 0.05 for peak calculations. The cut-off for end-points titres or the limit of positivity for AUC was defined as the value above the mean plus 3-times standard deviation of values obtained with the reactivity of respective proteins with pre-pandemic plasma samples of healthy donors. The fold-reduction in RBD-IgG cross-reactivity to VOCs was determined by dividing the median IgG-titer to wild-type RBD with the median IgG-titer obtained from VOC-RBD.

### SARS-CoV-2 Surrogate Virus Neutralization Test

The sVNT test was used to determine the neutralization potential of antibodies in blocking the RBD-ACE2 receptor interaction. The test was performed following the manufacturer’s instructions (Genscript). Briefly, 1:10 diluted plasma samples were incubated with RBD-HRP and the mixture was added on human ACE2 coated plate. The plate was developed using TMB substrate and read at 450 nm. The sample absorbance was inversely proportional to the titre of the anti-SARS-CoV-2 neutralizing antibodies. The percent neutralization was calculated using the formula: (1-OD value of sample/OD value of Negative Control) x 100%. The cut-off for the positive limit of SARS-CoV-2 neutralizing antibodies was determined by manufacturer and further validated using the plasma panel of pre-pandemic healthy controls ^4,20^.

### Detection of antigen-specific Memory B cells

To examine the quantity and phenotype of RBD-specific memory B cells and to test their cross-reactivity to the recent variants, polyclonal stimulated B cells were tested in the fluorospot assay utilizing RBD antigen from the ancestral strain of SARS-CoV-2 (B.1) and variants; alpha (B.1.1.7), beta (B.1.351), gamma (P.1), kappa (B.1.617.1) and delta (B.1.617.2). Briefly, cryopreserved PBMCs were thawed and polyclonally stimulated with R848 and IL-2 for 5 days. The stimulated cells were then captured on the Fluorospot plate (Mabtech) coated overnight with the respective RBD-antigen at 5µg/mL. For measuring total memory B cells of each isotype, wells were coated with anti-human IgG (MT91/145), anti-human IgM (MT11/12), and anti-human IgA (MT57) (Mabtech) at the concentration of 15µg/mL. Plates were washed, blocked and seeded with the 0.5×10^6^ stimulated PBMCs per well for antigen-specific analysis and 20,000 cells for total B cell analysis followed by incubation at 37°C for 8 hours. For detection of antibody secreting cell (ASC) spots corresponding to IgG^+^, IgM^+^ and IgA^+^ isotype, the plate was developed using detection monoclonal anti-human antibodies; IgG-550 (MT78/145), IgM-640 (MT22) and IgA-490 (MT20), diluted 1:500 in PBS containing 0.5% FBS for 2 hours in dark at room temperature. Plates were added with fluorescence enhancer (Mabtech), washed and spots were detected using AID vSpot Spectrum Elispot/Fluorospot reader system and analysed by AID Elispot software version 7.x. As no spots were detected in wells without the antigen, presence of a spot >1 in the antigen-coated well was considered as a positive response. ASC counts were normalized to ASCs per million of PBMCs for all analyses. The frequency of antigen-specific memory B cells was expressed as the percentage of total B cells representing respective isotype. The median frequency of B cells specific to wild-type RBD was divided by the median frequency of B cells reacting to the VOC-RBD and depicted as the fold-reduction in frequency for respective variant.

### Quantification of antigen-specific CD4^+^ T cells and cTfh cells

Antigen-specific CD4^+^ T cell were measured using the Activation-induced markers (AIM) assay, as described ^4,11,29^. Briefly, SARS-CoV-2-specific CD4^+^ T cells quantified as a percentage population of AIM^+^ (OX40^+^CD137^+^) CD4^+^ T cells after stimulation of PBMCs with overlapping peptide megapools (MPs) at a concentration of 1 μg/ml in AIM-V media (Gibco™, USA) for 24 hours at 37°C. The MPs consist of 15-mer peptides overlapping by 10 amino acids covering the Spike (S) or Nucleocapsid protein (NP) of ancestral SARS-CoV-2 (B.1; GenBank: MN_908947) and MPs covering Spike (S) protein of VOCs; Alpha (B.1.1.7; GISAID: EPI_ISL_601443), Beta (B.1.351; GIASID: EPI_ISL_660629) and Delta (B.1.617.2; GISAID: EPI_ISL_2020950). All the peptides were synthetized as crude material (TC Peptide Lab, USA), resuspended in DMSO, pooled and sequentially lyophilized as previously reported ^11,29^. An equimolar concentration of DMSO was used as a negative control. SEB at 0.1 µg/ml and Cytomegalovirus (CMV) specific MP were used as the positive stimulation control. After the stimulation period, cells were washed with FACS buffer (2% FBS in PBS) and surface stained with monoclonal antibody cocktail for 1 hour at 4° C in the dark; CD20, CD14, CD16, CD8a and fixable-viability dye coupled with APC eflour 780 in the dump channel, CD4-AlexaFluor 700 (RPA-T4), OX40-FITC (Ber-ACT35), CD137 PE Dazzle 594(4B4-1), CD45RA Brilliant Violet 785 (HI100) and CCR7 Alexa Fluor 647 (G043H7). The circulating Tfh (cTfh) cells were quantified by measuring the frequency of CXCR5^+^PD1^+/-^ cells in the AIM^+^CD4^+^ T cells obtained after stimulation with Spike and Nucleoprotein MPs. The frequency of cTfh cells was expressed as the percentage of total CD4^+^ T cells. For cTfh analyses, in addition to the AIM markers, cells were also stained with CXCR5-Brilliant Violet 421 (J252D4), PD1-PE (EH12.2H7), CXCR3-Brilliant Violet 711 (G025H7) and CCR6-PE/Cy7 (G034E3). For all analyses, after staining, cells were washed and fixed with freshly prepared 1% Paraformaldehyde (Sigma Aldrich) followed by acquiring using BD LSRFortessa X-20 flow cytometer (BD Biosciences) and subsequent data analysis by FlowJo 10.7.1. The antigen-specific CD4^+^ T cells were measured as the data subtracted from DMSO conditions as the background. The positive response in the AIM assay was defined by setting up the limit of positivity above the median plus two-times of standard deviation of negative controls (pre-pandemic PBMCs) stimulated with spike Peptide_MP_^4^. The frequency of responders to SARS-CoV-2 spike Peptide_MP_ stimulation was determined by applying the Fischer’s exact test on the AIM^+^ and AIM^-^ cells in DMSO and Peptide_MP_ stimulated conditions.

### Bead-Based Multiplex Cytokine Immunoassay

The cytokine levels were measured in the culture supernatants of PBMCs stimulated for 24 hour with SARS-CoV-2 spike Peptide_MP_. The cell free supernatants were stored at -80°C until thawed for the quantifications of secreted factors. The T-cell specific secreted cytokines were quantified using the Bio-Plex human cytokine screening panel (17-Plex, Bio-Rad) on a Luminex 200 multiplex suspension array system, following the manufacturer’s instruction. The positive cytokine response was defined as the response above the lowest limit of quantification and significance of p>0.05 between unstimulated and spike Peptide_MP_ stimulated condition.

### Qualitative assessment of SARS-CoV-2-specific memory CD4^+^ T cells

The B cell help quality of antigen-specific memory CD4^+^ T cells in vaccination and infection was assessed in the autologous T:B co-culture assay, as we described recently ^47^. Briefly, cryopreserved PBMCs were revived as mentioned previously with ≥90% cellular viability and surface stained with following monoclonal antibodies for 15 minutes at 4 °C: fixable viability dye efluor 506, anti-CD4 AF700 (RPA-T4), anti-CD45RO FITC (UCHL1), anti-CD14 PE (MΦP9) and anti-CD20 PE-Cy7 (2H7). Memory CD4^+^ T cells sorted as live CD14^-^CD20^-^ CD4^+^CD45RO^+^ cells. Live CD20^+^CD14^-^ cells and CD14^+^CD20^-^ cells were sorted as B cells and monocytes, respectively. Cells were sorted on a BD FACSAria Fusion flow cytometer (BD Biosciences) at low flow rate using a 70μm nozzle. Autologous memory CD4^+^ T cells and CD20^+^ B were co-cultured in 1:1 ratio (6×10^4^ cells/per well) in presence of monocytes (3×10^4^ cells) in AIM-V serum free medium (Gibco) in 96-well plates (Nunc, Thermo). Cells were kept either unstimulated (no exogenous stimulation) or stimulated with SARS-CoV-2 full length spike protein (CoV-S protein, 10μg/mL) (Native Antigen, UK) for 9 days. After 9 days, enumeration of plasma cells as CD20^lo^CD38^hi^ in CD27^+^CD19^+^ B cells was performed by flow cytometry. Cells were surface stained with following antibodies for 40 minutes at 4 °C: fixable viability dye efluor 506 (eBioscience), anti-CD3 APC-Cy7 (HIT3a), anti-CD19 BV786 (HIB19), anti-CD27 PE-Dazzle594 (M-T271), anti-CD20 PE-Cy7 (2H7) and anti-CD38 PE-Cy5 (HIT2). After surface staining cells were washed with FACS buffer (2% FBS in PBS) and resuspended in FACS buffer. All the samples were acquired on BD LSRFortessa X-20 flow cytometer (BD Biosciences). Data were analyzed using FlowJo 10.7.1.

### IL-21 ELISpot

IL-21 secreting T cells were quantified using human IL-21 ELISpot (3540; Mabtech) in the cells obtained from T:B co-cultures, after 9 Days of stimulation with spike protein. Briefly, multiscreen ELISpot plate (MSIPS4510; Millipore) was coated with mAb MT216G (10μg/mL, Mabtech) for overnight at 4°C. Plate was washed and blocked with 10% FBS for at least 30 minutes at room temperature. After blocking of the wells, cells from co-cultures were seeded and restimulated with SARS-CoV-2 spike protein (2μg/mL) for 24 hours at 37°C. Next day, plate was washed rigorously with PBS and developed by incubating with detection antibody mAb MT21.3m-biotin (Mabtech) for 2 hours followed by the addition of Streptavidin-ALP and BCIP/NBT substrate. The plate was dried and read on AID vSpot Spectrum ELISpot/Fluorospot reader system. IL-21 SFCs were captured and enumerated using AID ELISpot software version 7.x.

### IFN-γ Fluorospot

To enumerate IFN-γ secreting cells, Fluorospot assay was performed according to manufacturer’s protocol (Mabtech, Sweden). Low fluorescent IPFL 96 well plate (Mabtech, Sweden) was coated with 15 μg/ml of purified mAb IFN-γ (clone 1-D1K, Mabtech, Sweden) for overnight at 4°C. After incubation, plate was washed, blocked and seeded with 250×10^3^ PBMCs followed by the stimulation with SARS-CoV-2 spike Peptide_MP_ at 1µg/ml for 24 hour. Equimolar DMSO was used as negative control and SEB at 0.1 µg/ml was used as positive stimulation control. After incubation, IFN-γ spots were detected using BAM conjugated monoclonal antibody against IFN-γ (7-B6-1-BAM, Mabtech, Sweden). The plate was developed by incubating with the anti-BAM 490 (Mabtech, Sweden) and addition of fluorescent enhancer (Mabtech, Sweden). The plate was dried and spots were read using AID vSpot Spectrum ELISpot/Fluorospot reader system and quantified using AID ELISpot software version 7.x.

### Statistical analyses

In all the experiments, data are either represented as median, or as mentioned in the figure legends. The significance of the differences between the groups was analysed with the two-sided Mann-Whitney test, Fischer’s exact test, Wilcoxon matched-pairs signed rank test, two-way ANOVA or as specified in the figure legends. P values <0.05 were considered statistically significant. Statistical analyses and data visualization were performed with the GraphPad Prism software version v8.4.3.

## Data Availability

All data are available in the main text or the extended data

## Acknowledgments

We are thankful to all the patients and vaccinees for generous support in this study. This research has been conducted with the contribution of NCR Biotech Science Cluster BIOREPOSITORY supported by DBT. We thank all the clinical, laboratory, and data management staff of all partnering medical centres for their contributions to this work. At the National Institute of Immunology, we thank Mr Rabindra Prasad at the Flow cytometry core facility. Some schematics were created with Biorender.com. This work was financially supported by the Science and Engineering Research Board, DST grant (IPA/2020/000077) to NG, Department of Biotechnology (DBT) grant (BT/PR30223/MED/2018) to NG and the core grant of National Institute of Immunology. COVID-19 Bioresource grant (BT/PR40401/BIOBANK/03.2020) to Translational Health Science and Technology Institute. Further support provided in part with Federal funds from the National Institute of Allergy and Infectious Diseases, National Institutes of Health, Department of Health and Human Services, under Contract No. 75N93021C00016 to A.S. and Contract No. 75N9301900065 to A.S, D.W.

## Author contributions

N.G., conceptualized, supervised the study and wrote the original draft, R.V, A.A, S.N.J, A.R.P, Sh.S., S.P., B.N., A.K., H.A.P, T.S., carried out the investigation, B.P.J., S.S., P.K., N.W., P.C., S.K., P.S., N.S., J.T., A.K.P., As.S., R.T., S.B., supervised the enrolment and sample collection, S.B., A.G., D.W., P.K.G., A.S., provided the resources, N.G., S.B., A.S., funding acquisition. All authors edited the manuscript and approved the final submission.

