## Extended Data for "Inactivated virus vaccine BBV152/Covaxin elicits robust cellular immune memory to SARS-CoV-2 and variants of concern"

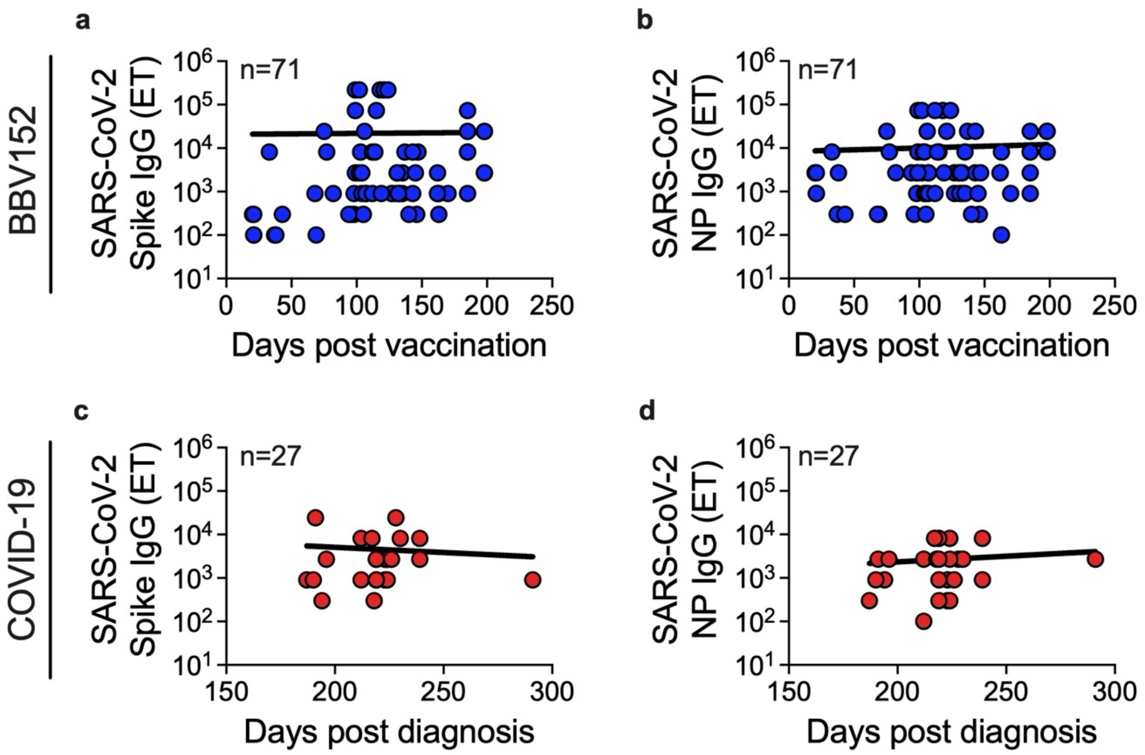


**Extended data Fig. 1. BBV152-elicited antibodies persisted at least up to 6 months after complete vaccination.** The antibody response was measured as end-point titers in BBV152 vaccinated (n=71) and COVID-19 recovered subjects (n=27), as mentioned in the methods and legends of Fig. 1. Persistence of (**a**) anti-spike and (**b**) anti-nucleoprotein (NP) IgG over the days post vaccination, and (**c**) anti-spike and (**d**) anti-nucleoprotein IgG over the days post-diagnosis of COVID-19, in BBV152 vaccination and recovery from mild COVID-19, respectively. Statistics by non-linear regression analysis.

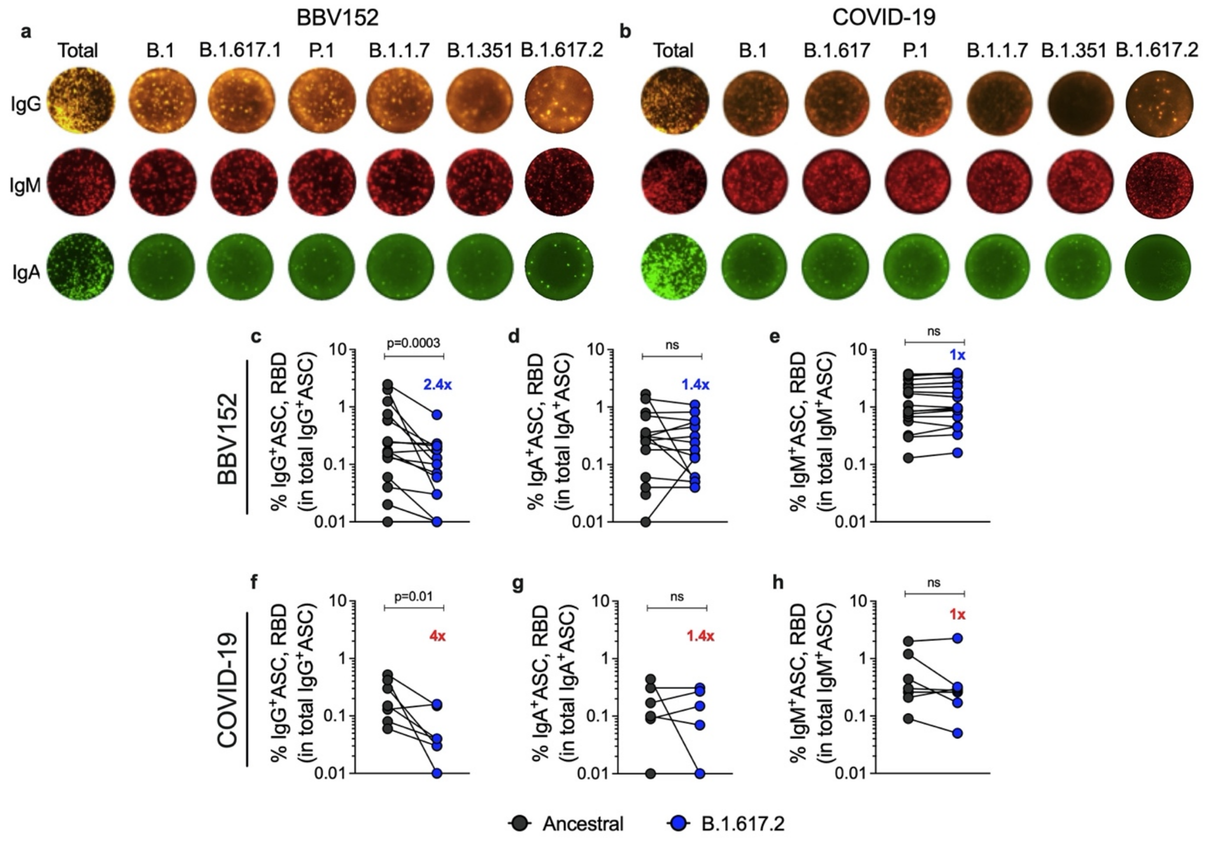


**Extended data Fig. 2. BBV152-induced memory B cells robustly responded to B.1.617.2 (Delta) variant.** Frequency and isotype distribution of memory B cells specific to RBD of ancestral SARS-CoV-2 and Delta variant was measured in individuals vaccinated with BBV152 ("BBV152", n=17) and individuals recovered from mild COVID-19 ("COVID-19", n=7). The RBD-specific memory B cells were enumerated in the R848+IL2 stimulated B cells utilizing the antigen-specific ELIspot analysis. Representative ELIspot images of $IgG^+$, $IgM^+$ and $IgA^+$ memory B cells against RBD of ancestral SARS-CoV-2 and its variants in (**a**) vaccinated and (**b**) recovered COVID-19 subjects. "Total" represent the frequency of total B cells captured using anti-IgG, -IgM or -IgA antibody. Proportion of SARS-CoV-2-RBD-specific antibody secreting cells (ASCs) was measured by calculating the percentage of antigen-specific B cells in the total corresponding isotype of ASCs in 1 million PBMCs. Comparison of the frequency of ancestral SARS-CoV-2 RBD-specific memory B cells and their reactivity to RBD protein of Delta variant for (**c**) $IgG^+$ ASCs (**d**) $IgA^+$ ASCs and (**e**) $IgM^+$ ASCs in BBV152 vaccination, and for (**f**) $IgG^+$ ASCs (**g**) $IgA^+$ ASCs and (**h**) $IgM^+$ ASCs in recovery from mild COVID-19. Quantitative reduction in RBD-reactive B cells against Delta variant over the RBD-specific B cells to ancestral virus is depicted in top of graph. Statistics by (**c-h**) Wilcoxon signed-rank test. ns: non-significant.

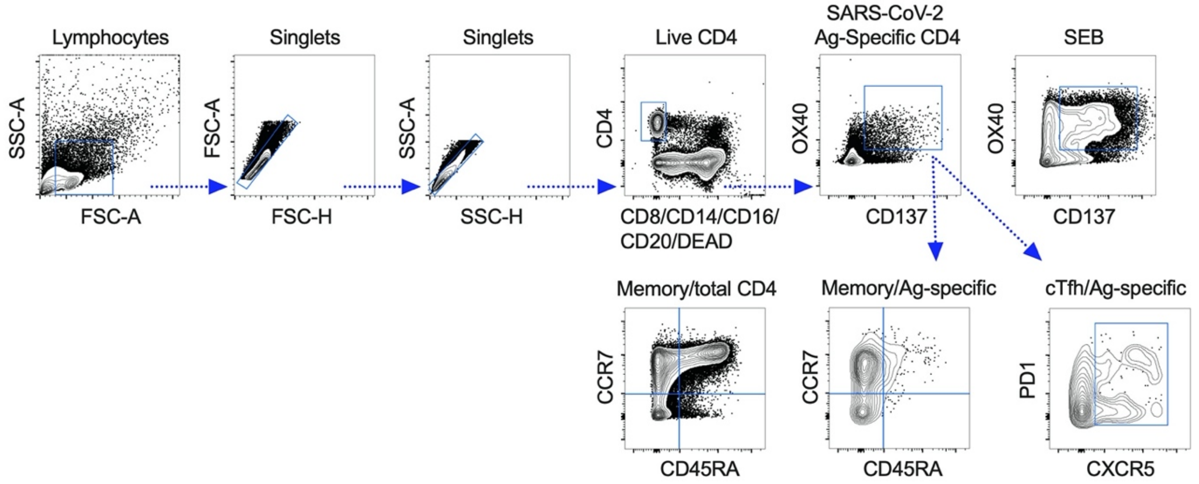


**Extended data Fig. 3. Flow cytometry Gating Strategy**. The gating strategy for quantitation of the antigen-specific CD4$^+$ T cells as determined by OX40$^+$CD137$^+$ (AIM$^+$) cells, total and antigen-specific memory CD4$^+$ T-cell subsets, and circulating total and SARS-CoV-2-specific follicular helper T (Tfh) cells in the PBMCs derived from individuals vaccinated with BBV152 and individuals recovered from mild COVID-19.

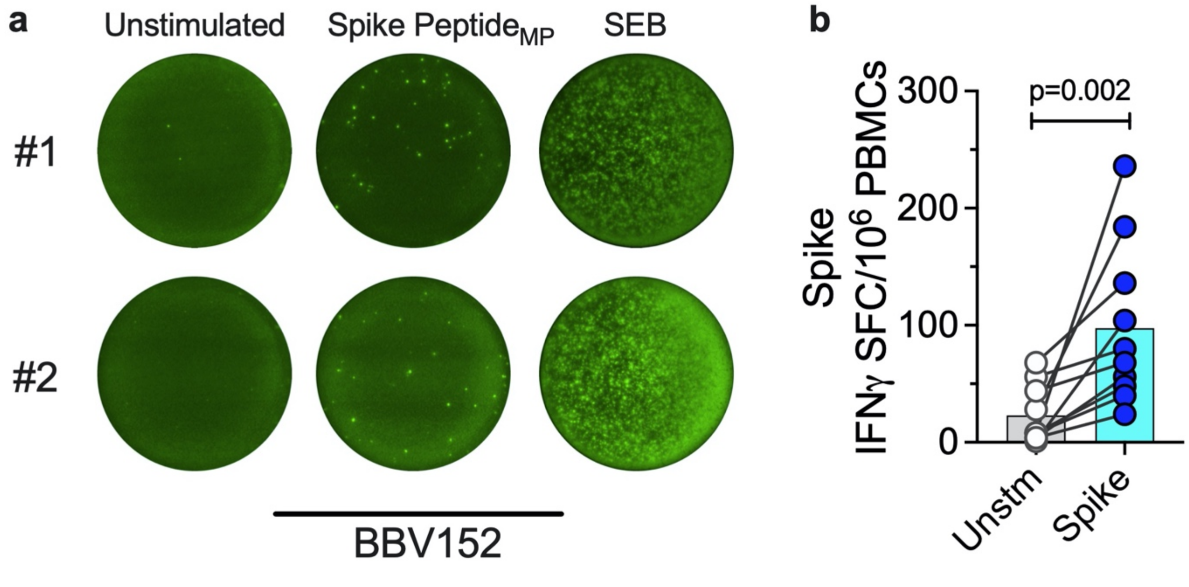


**Extended data Fig. 4. BBV152 vaccination induces IFN-γ expressing cells**. The IFN-γ spot forming cells were measured in the PBMCs derived from individuals with BBV152 vaccination (n=10) after stimulation with overlapping peptide megapools (MP) spanning spike protein of SARS-CoV-2 in the IFN- γ Fluorospot assay. (**a**) Representative images of IFN-γ spot forming cells (SFCs) in PBMCs from two vaccinated donors, stimulated with DMSO (Unstim), spike peptide$_{MP}$ (spike), and Staphylococcal Enterotoxin B (SEB) as a positive control. (**b**) The magnitude of IFN-γ spot forming cells (SFCs) per million PBMCs was compared between unstimulated and the spike peptide$_{MP}$ stimulated conditions. Statistics by Wilcoxon signed-rank test.

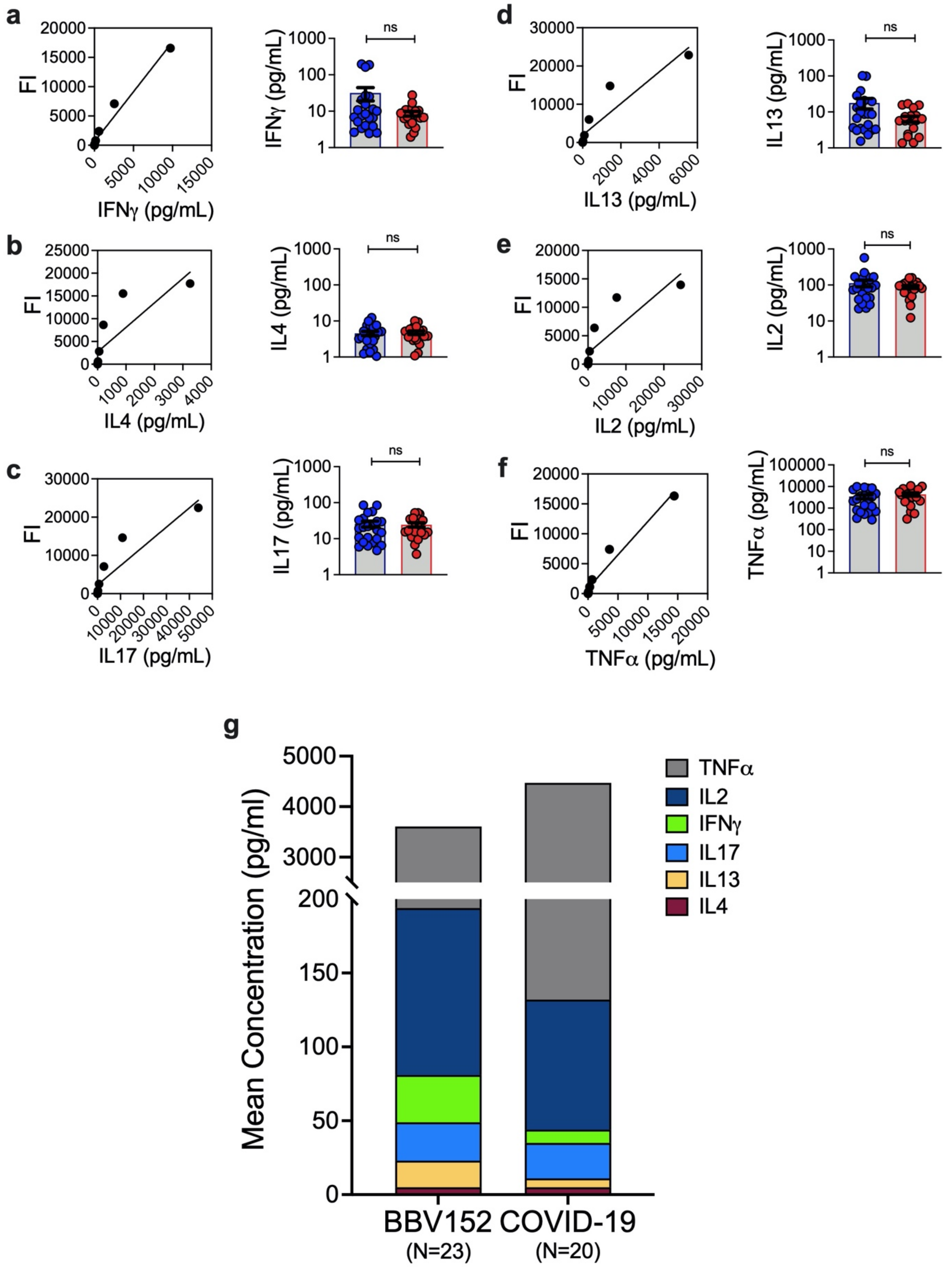


**Extended data Fig. 5. BBV152 vaccination generates multifunctional spike-specific CD4**$^{+}$ **T-cell response with the polarization towards Th1-cell phenotype**. Cytokine levels were measured by multiplex bead-based immunoassay in the cell culture supernatants of PBMCs stimulated or not with spike peptide$_{MP}$ for 24h, and compared between BBV152 vaccination (n=23) and recovery from mild COVID-19 (n=20). The standard curve depicting the fluorescence intensity (FI) of cytokine-standard and the adjacent bar graph showing the concentration (pg/ml) of T-cell specific cytokines in vaccination and infection for (**a**) IFN-γ (**b**) IL4 (**c**) IL17 (**d**) IL13 (**e**) IL2 and (**f**) TNF-α. (**g**) Stacked graph showing the comparison of mean concentration of each cytokine in BBV152 vaccination and recovery from mild COVID-19. The data denotes the mean±s.e.m. The positive response was determined as the response over Lower Limit of Quantification (LLOQ) and statistical significance in spike $peptide_{MP}$ stimulation over unstimulated conditions. Statistics by two-tail Mann-Whitney test. ns: non-significant.

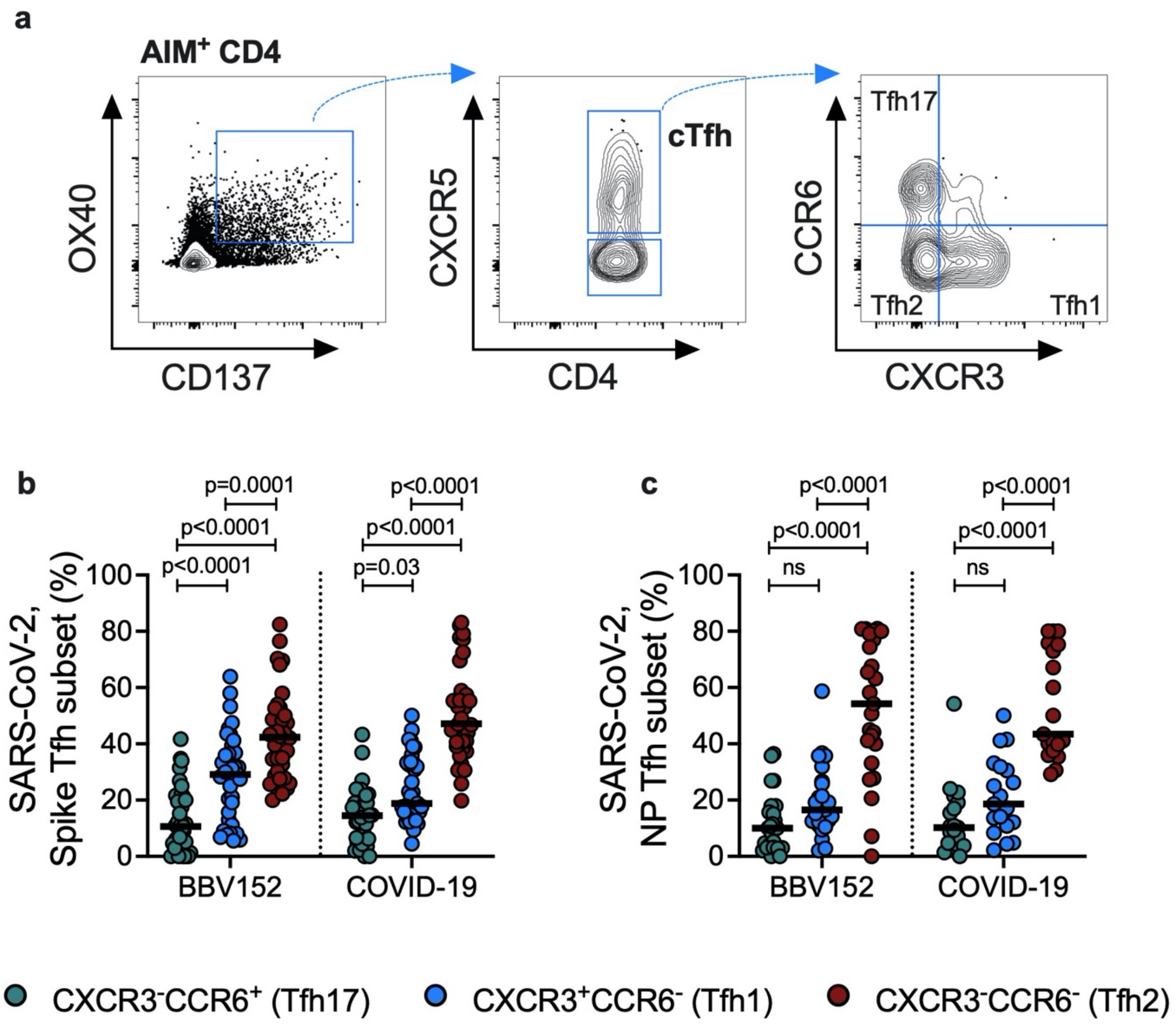


**Extended data Fig. 6. BBV152-induced circulating Tfh cells comprised of both Tfh1 and Tfh2 phenotype.** The functional heterogeneity of spike and nucleoprotein (NP) specific circulating follicular helper T (cTfh) cells was determined based on the expression of CXCR3 and CCR6 in total Tfh cells ($CXCR5^+$) among the $AIM^+$ antigen-specific $CD4^+$ T cells. The proportion of Tfh subsets was measured in the SARS-CoV-2 specific cTfh cells induced in response to BBV152 vaccination or mild COVID-19. (**a**) Representative gating strategy for measuring the Tfh heterogeneity in SARS-CoV-2-specific $CD4^+$ T cells. Proportion of Tfh17 ($CXCR3^-CCR6^+$), Tfh1 ($CXCR3^+CCR6^-$), and Tfh2 ($CXCR3^-CCR6^-$) subsets among (**b**) spike-specific cTfh and (**c**) nucleoprotein-specific cTfh cells in BBV152 (spike: n=34; NP: n=25) and COVID-19 (spike: n=37; NP: n=19) groups. Black bars indicate median. Statistics by 2way ANOVA followed by Tukey's multiple comparisons. ns: non-significant.
